# Associations of behavioral problems with white matter circuits connecting to the frontal lobes in school-aged children born at term and preterm

**DOI:** 10.1101/2023.11.08.23298268

**Authors:** Machiko Hosoki, Margarita Alethea Eidsness, Lisa Bruckert, Katherine E. Travis, Heidi M Feldmana

## Abstract

**Introduction:** This study investigated whether behavioral problems in children were associated with fractional anisotropy (FA) of white matter tracts connecting from other brain regions to right and left frontal lobes. We considered internalizing and externalizing behavioral problems separately and contrasted patterns of associations in children born at term and very preterm.

**Methods:** Parents completed the Child Behavior Checklist/6-18 questionnaire to quantify behavioral problems when their children were age 8 years (N=36 FT and 37 PT). Diffusion magnetic resonance scans were collected at the same age and analyzed using probabilistic tractography. We used multiple linear regression to investigate the strength of association between age-adjusted T-scores of internalizing and externalizing problems and mean fractional anisotropy (mean-FA) of right and left uncinate, arcuate, and anterior thalamic radiations, controlling for birth group and sex.

**Results:** Regression models predicting internalizing T-scores from mean-FA found significant group-by-tract interactions for the left and right arcuate and right uncinate. Internalizing scores were negatively associated with mean-FA of left and right arcuate only in children born at term (*p*_left AF_ =0.01, *p*_right AF_ =0.01). Regression models predicting externalizing T-scores from mean-FA found significant group-by-tract interactions for the left arcuate and right uncinate. Externalizing scores were negatively associated with mean-FA of right uncinate in children born at term (*p*_right UF_ =0.01) and positively associated in children born preterm (*p*_right UF preterm_ =0.01). Other models were not significant.

**Conclusions:** In this sample of children with scores for behavioral problems across the full range, internalizing and externalizing behavioral problems were negatively associated with mean-FA of white matter tracts connecting to frontal lobes in children born at term; externalizing behavioral problems were positively associated with mean-FA of the right uncinate in children born preterm. The different associations by birth group suggest that the neurobiology of behavioral problems differs in the two birth groups.

**’Highlights’:**

⍰ Internalizing problems were negatively associated with arcuate FA in term children.

- Externalizing problems negatively associated with right uncinate FA in term children.
- Externalizing problems were positively associated with right uncinate FA in preterm.

## Introduction

Mental health conditions in children and youth have become increasingly recognized as critical public health issues (Kessler et al., 2009; Patel et al., 2007; Whiteford et al., 2013). Parent-reported behavioral problems, assessed on standardized questionnaires, may be precursors to or early indicators of mental health problems and may impact long-term academic success and social competence (Liu et al., 2011). During childhood, behavioral problems are typically divided into two categories: internalizing and externalizing problems (Liu et al., 2011; McMahon, 1994; Montoya-Castilla et al., 2018; Ollendick and King, 1994). Internalizing problems indicate internalized expressions of pain or distress, such as anxiety, fear, shyness, somatic complaints, withdrawal, and depression (Bongers et al., 2003; Ollendick and King, 1994; Wei and Lü, 2023). Externalizing problems represent outward expressions of distress, such as defiance, aggression, destructiveness, and conduct problems (McMahon, 1994; Wei and Lü, 2023). Behavioral problems are highly prevalent. The National Survey of Children’s Health reported that among children between the ages of 3 to 17 years, the prevalence rate was 3.2% for depression, 7.1% for anxiety, and 7.4% for conduct problems (Ghandour et al., 2019). Internalizing and externalizing problems often show a high degree of covariance (Hatoum et al., 2018a). Behavioral problems are likely caused by a convergence of biological and environmental factors (Hatoum et al., 2018b). Understanding these factors may provide clues to prevention or treatment strategies to reduce their prevalence or severity. In this study, we investigated a possible neurobiological factor associated with internalizing and externalizing behavioral problems in children, focusing on the microstructure of white matter circuitry in the brain. We compared the pattern of associations in two distinct populations, children born at term and children born preterm, because children born preterm are at higher risk for behavioral problems and disturbances of white matter development.

The literature has identified the frontal lobe as a brain region that modulates emotional and social processing and thereby may be involved in the development, presence, or intensity of behavioral problems in children. For example, the orbitofrontal cortex (OFC), rostral middle frontal cortex, and the anterior cingulate cortex (ACC) have been consistently associated with internalizing symptoms (Blok et al., 2022). The OFC, ACC, amygdala, hippocampus, and striatum have been linked to externalizing symptoms (Blok et al., 2022).

A growing literature reports that alterations in white matter can be observed in mental health conditions. These findings may indicate primary disturbances with a causative role or secondary effects of variation in impulse propagation (Fields, 2008). Assessment of white matter circuits might provide clues to causes or to treatments of these mental health conditions. Several white matter tracts connect these regions of the frontal lobe to other brain regions. In this study we focused on three that have been implicated in mental health conditions and can be reliably reconstructed in children using diffusion magnetic resonance imaging (dMRI). The anterior thalamic radiation (ATR) connects the dorsolateral prefrontal cortex (DLPFC) with the thalamus through the anterior limb of the internal capsule (Deng et al., 2018). The uncinate fasciculus (UF) connects the orbital, medial, and lateral PFC with rostral temporal areas, including the anterior temporal lobe, parahippocampal gyrus, entorhinal and perirhinal cortex, and amygdala (Koch et al., 2017). The arcuate fasciculus (AF) connects the ventrolateral and dorsolateral part of the PFC with the posterior parieto-temporal regions (Nakajima et al., 2018). Individual differences in microstructure or white matter pathways are often assessed using fractional anisotropy (FA), which measures the directionality of axons within a white matter tract.

Among clinical populations with diagnosed mental health conditions or observed differences in behavioral characteristics, many research studies have found associations between a range of behavioral symptoms and the microstructure of three white matter tracts connecting to the frontal lobe, the ATR, UF and AF. Adults and adolescents with major depressive disorder exhibit abnormal diffusion measures, mainly in the DLPFC part of the left ATR (Bessette et al., 2014; Hasan et al., 2008; Zhu et al., 2011). Among infants with a history of neglect and maltreatment, decreased FA of UF was associated with elevated self-reported internalizing problems (Hanson et al., 2015). Studies have also found positive correlation between mean diffusivity (MD) and reduced FA in the right UF, exacerbating anxiety scores (Ho et al., 2017; Koch et al., 2017). Other studies have found a correlation between UF and symptoms such as flattened affect and lack of social engagement (Craig et al., 2009; Motzkin et al., 2011). Patients with bipolar disorder (BD) exhibited lower FA in the left AF anterior segment than did healthy controls (Sarrazin et al., 2014). In veterans, decreased FA of part of AF was associated with aggression and anger outbursts (David et al., 2020). Based on this review, features of the ATR, UF and AF are likely candidates to assess whether white matter circuits to the frontal lobe might be associated with behavioral problems in children.

Preterm birth is associated with an increased risk for impaired neurobehavioral development (Batalle et al., 2017; Kallankari et al., 2021; Vanes et al., 2022). Perinatal factors such as low birth weight and gestational age have been associated with an increased risk of developing mental health disorders in early adolescence and persisting into young adulthood (Anderson et al., 2021; Johnson and Marlow, 2011; Vanes et al., 2022). Preterm-born children, particularly those classified as very preterm, with a gestational age less than 32 weeks, have consistently shown increases in externalizing and internalizing behaviors in childhood (Bhutta et al., 2002). Thus, this population is clinically at risk for developing internalizing and externalizing problems compared to their age-equivalent peers born at term.

Very preterm birth has been associated with long-term changes in the microstructure of white matter. Studies on extreme preterm infants demonstrate altered structural connectivity of cortico-basal ganglia-thalamo-cortical loop connections (CBTCL) (Fischi-Gómez et al., 2015). Prematurity-associated alterations in white matter were found in cortical and subcortical structures, affecting projection, commissural, and association fibers (Pandit et al., 2014). Additionally, adults with a history of very preterm birth have altered white matter properties (Kelly et al., 2023). However, studies differ as to whether FA values are increased or decreased when comparing individuals born preterm with peers born at term.(Kelly et al., 2023; Travis et al., 2015).

Research on the association of behavioral problems comparing children born at term and preterm and white matter circuitry to frontal lobe is limited. In a study using tract-based spatial statistics among school-aged children born at term and preterm, negative correlations between parent-reported Child Behavior Checklist (CBCL) internalizing problem scores and FA of tracts coursing to the frontal lobes, including forceps minor, inferior fronto-occipital fasciculus, and right superior longitudinal fasciculus were found only in the preterm group. In addition, the same study found negative correlations only among children born preterm between FA and parent-reported CBCL externalizing problems in the forceps major and inferior fronto-occipital fasciculus (Loe et al., 2013). In another tractography study using fixel-based analysis (FBA), Gilchrist et al. found significant associations of internalizing and externalizing problems with fiber density (FD), fiber-bundle cross-section (FC), and fiber density and cross-section (FDC) in several white matter pathways (Gilchrist et al., 2023) in 7-year-olds born both term and preterm. Significant associations were limited to the right UF in children born at term at age 13 years.

In this study, we assessed whether among school-aged children born at term and preterm, internalizing and/or externalizing problems would correlate with FA of three white matter tracts that connect to the frontal lobe, the ATR, UF and AF. We analyzed white matter circuitry using probabilistic diffusion magnetic resonance imaging (dMRI) tractography. We focused on these tracts because the literature has implicated them as associated with behavior problems and they can be reliably reconstructed in children (Bruckert et al., 2019). Based on the results of the study by Loe and colleagues (Loe et al., 2013), we anticipated significant negative associations of FA and behavior problems within the children born preterm. Based on the results of the FBA analysis by Gilchrist and colleagues (Gilchrist et al., 2023), we recognized that internalizing and externalizing behavioral problems might be associated with microstructural properties of the multiple tracts, particularly the UF, in both children born at term and preterm.

## Materials and Methods

### Participants

The participants were children born at term and preterm enrolled in a longitudinal study of reading skills and white matter characteristics (Bruckert et al., 2019; Dubner et al., 2020). Children were recruited from the San Francisco Bay Area from 2012 to 2015 and were followed for 2 years, from age 6 to 8 years. Preterm birth was defined as gestational age ≤ 32 weeks at birth and full-term birth was defined as gestational age ≥ 37 weeks or birth weight ≥ 2500 grams. We restricted enrollment to children born ≤ 32 weeks as children born preterm because very preterm birth increases the risk for white matter injury (Larroque et al., 2003) and because we anticipated they would have elevated behavioral problems (Aarnoudse-Moens et al., 2009). Exclusion criteria included hearing loss, visual impairment, history of neurological disorders, non-English speakers, and genetic disorders.

For this analysis, we included 8-year-old children whose parents completed the Child Behavior Checklist (CBCL), who underwent diffusion MRI, and whose scans were found to be free of motion artifacts and other technical problems, as described in previous studies of the sample (Bruckert et al., 2023; Hosoki et al., 2022). We decided to analyze the children at age 8-years because we would likely capture more behavioral problems at the older age than at age 6-years, given the nature and longitudinal emergence of behavioral problems (Scott et al., 2018). The final sample included 73 children, 36 children born at term (16 boys) and 37 children born preterm (20 boys).

The experimental protocol was approved by the Stanford University Institutional Review Board. Written consent was obtained from a parent or a legal guardian and written assent was obtained from the participant. Children were compensated for participation. We collected demographic characteristics, including child sex assigned at birth and birth group.

### Assessments conducted at age 8

Parents completed the parent-reported Child Behavior Checklist (CBCL/6-18), a questionnaire that indexes children’s behavioral problems (Achenbach and Edelbrock, 1983). The instrument generates broad-band and narrow-band scores. We focused on the main broad bands— internalizing and externalizing subscales—because generally broad band scores have greater accuracy than narrow band scores. Internalizing problems include narrow band scores, which are anxious/depressed behavior, withdrawn-depressed behavior, and somatic complaints. Externalizing problems include narrow band scores, which are aggressive behavior and rule-breaking behavior (Achenbach and Ruffle, 2000). We analyzed T-scores of internalizing and externalizing problems.

### Diffusion MRI acquisition and analysis

Brain MRI data were acquired at age 8 years using a 3T scanner (GE MR750 Discovery; GE Healthcare, Waukesha, WI, USA) with a 96-channel head coil. All children were scanned for research purposes, without the use of sedation. MRI data analyzed in the current study included i) a high resolution T1-weighted scan using a 3D fast-spoiled gradient (FSPGR) sequence (TR = 7.24 ms; TE = 2.78 ms; FOV = 230 mm × 230 mm; acquisition matrix = 256 × 256; 0.9 mm isotropic voxels; orientation = sagittal) and ii) a diffusion MRI scan using a dual-spin echo, echo-planar imaging sequence (96 directions, b = 2500 s/mm^2^, 3 b =0 volumes, voxel size = 0.8549 × 0.8549 × 2 mm^3^,TR = 8300 ms, TE = 83.1 ms).

### Diffusion MRI analysis

MRI data were managed and analyzed using a neuroinformatics platform (Flywheel.io) that guarantees data provenance, implements reproducible computational methods, and facilitates data sharing. The diffusion MRI analysis pipeline consisted of diffusion MRI preprocessing, whole-brain tractography, and tract segmentation. The steps were described in detail by Lerma-Usabiaga et al. (Lerma-Usabiaga et al., 2019) and Liu et al. (Liu et al., 2022) and are briefly summarized below.

### Diffusion MRI preprocessing

Diffusion MRI data were preprocessed using a combination of tools from MRtrix3 (github.com/MRtrix3/mrtrix3), FSL (https://fsl.fmrib.ox.ac.uk/), and mrDiffusion part of VISTASOFT (http://github.com/vistalab/vistasoft) as implemented in the Reproducible Tract Profiles (RTP) pipeline (Lerma-Usabiaga et al., 2019). In brief, the steps of preprocessing included (i) denoising the data using principal component analysis (Veraart et al., 2016b, 2016a) and Gibbs ringing correction (Kellner et al., 2016), (ii) applied FSL’s eddy current correction (Andersson and Sotiropoulos, 2016), (iii) resampling the data to 2 x 2 x 2 mm^3^ isotropic voxels, and (iv) aligning the diffusion data to the average of the non-diffusion-weighted volumes, which were aligned to the child’s high-resolution anatomical image using rigid body transformation.

### Diffusion MRI tractography

The preprocessed diffusion MRI data served as the input for whole-brain tractography and tract segmentation. We used the constrained spherical deconvolution model (CSD, Tournier et al., 2007) with eight spherical harmonics (lmax = 8) to calculate fiber orientation distributions (FOD) for each voxel. These FODs were used for diffusion MRI tractography, which consisted of the following steps: (i) Ensemble Tractography (Takemura et al., 2016) to estimate the whole-brain white matter connectome. MRtrix3 (Batalle et al., 2017) was used to generate three candidate connectomes that varied in their minimum angle parameters (50°, 30°, 10°). For each candidate connectome, a probabilistic tracking algorithm (iFOD2) was used with a step size of 1 mm, a minimum length of 10 mm, a maximum length of 200 mm, and an FOD stopping criterion of 0.04. (ii) Spherical-deconvolution Informed Filtering of Tractograms (SIFT) to improve the quantitative nature of the ensemble connectome by filtering the data such that the streamline densities match the FOD lobe integral. The resulting ensemble connectome retained 500,000 streamlines. (iii) Concatenation of the three candidate connectomes into one ensemble connectome. (iv) Automated Fiber Quantification (AFQ) (Yeatman et al., 2012) was to segment the resulting whole-brain connectome of each child into our tracts of interest: bilateral ATR, AF and UF. Tract segmentation was done using a way-point Region Of Interest (ROI) approach as described by Wakana et al. (Wakana et al., 2007). ROIs were transferred from a standard template to the native space of the participant so that tracking was done in native space. The tracts were refined by removing streamlines that were more than 5 standard deviations away from the core of the tract or that were more than 4 standard deviations above the mean streamline length (Yeatman et al., 2012). While CSD was used to model the fibers (as it can discern crossing fibers), the diffusion tensor model (DTI) was used to calculate FA, a commonly used diffusion metric.

### Statistical analysis

All statistical analyses were conducted using R studio with statistical significance set at p<0.05. We conducted chi-squared tests to compare the proportions of boys by birth group. We used independent samples t-test to compare internalizing problem T-scores, externalizing problem T-scores, and mean-FA of cerebral tracts by birth group. We used Welch t-test if there was unequal variance. To confirm we should analyze internalizing and externalizing problems separately, we ran a Pearson correlation to determine the magnitude of correlation between internalizing problem T-scores and externalizing problem T-scores, anticipating a low or moderate level of association.

We conducted a series of hierarchical linear regression models to assess the contribution of mean-tract FA of each of the cerebral tracts to CBCL internalizing problem T-scores and to externalizing problem T-scores at age 8. We adjusted for sex if there was a significant difference in sex by birth group. Birth group and sex were dummy coded. First, we assessed the contribution of sex and birth group to internalizing and externalizing problem T-scores. We then incorporated the contribution of the mean-tract FA of the cerebral tracts to the outcomes (main effect of tract). We computed R^2^ and compared R^2^ change between the model that investigated the main effect of the mean-FA of cerebral tracts to the model that only included covariates, to determine the contribution of mean-tract FA of the tract to the outcome. We next evaluated the interaction term to determine if birth-group moderated the association between behavioral problems and mean-tract FA of the cerebral tract. We then compared R^2^ change between the model with the interaction term to the model that investigated the main effect of mean-tract FA of the cerebral tract. We analyzed slopes of the association when the interaction term was significant.

To determine if associations of tract and behavior problems were specific to internalizing or externalizing problems, we further evaluated those cerebral tracts in which we found a significant association between both internalizing problem T-scores and externalizing problem T-scores. We ran two regression models in which we considered the contribution of the mean-tract FA of the cerebral tract to internalizing problem T-scores while adjusting for covariates and externalizing problem T-scores and vice versa. If there was a cerebral tract that demonstrated association between internalizing problem T-scores and externalizing problem T-scores respectively and the degree of association differed by birth group, we ran another model; we assessed the contribution of the mean-tract FA of the cerebral tract to externalizing problem T-scores by birth group, while adjusting for covariates and internalizing problem T-scores.

## Results

### Participant Characteristics

Characteristics of participants are presented in Table 1. There were birth group differences in sex; therefore, sex was included as a covariate in the linear regression models. There was no birth group difference in scores on the behavioral problem scale. Comparing FA of the different tracts, difference in left UF mean-tract FA by birth group was the only statistically result. The Pearson correlation coefficient of internalizing problem T-score and externalizing problem T-score was 0.47, confirming a moderate but not strong correlation and justifying separate analyses for internalizing problems and externalizing problems.

**Table 1.** Characteristics of the sample in terms of sex distribution, T-scores for internalizing and externalizing problems and mean-tract Fractional Anisotropy (FA) for the assessed tracts.

| Variable | Term (n=36) M(SD)<br>or n(%) | Preterm (n=37)<br>M(SD) or n(%) | t or X<sup>2</sup> | p-value |
| --- | --- | --- | --- | --- |
| Boys (%) | 16 (44%) | 20 (54 %) | X<sup>2</sup>(1)= 3.96 | **0.047** |
| Behavior scores |  |  |  |  |
| CBCL<sup>1</sup> Internalizing<br>problem T scores<br>M(SD) | 47.2 (11.0) | 50.6 (9.6) | t(71)= -1.40 | 0.165 |
| CBCL<sup>1</sup> Externalizing<br>problem T scores | 46.2 (11.2) | 46.67 (9.1) | t(62.6)= -0.20 | 0.842 |
| Mean-tract FA<sup>2</sup> of<br>Tracts |  |  |  |  |
| Left ATR<sup>3</sup> | 0.45 (0.04) | 0.43 (0.04) | t(71)= 1.80 | 0.076 |
| Right ATR<sup>3</sup> | 0.43 (0.05) | 0.42 (0.04) | t(71)= 1.60 | 0.114 |
| Left AF<sup>4</sup> | 0.48 (0.03) | 0.47 (0.03) | t(71)= 1.05 | 0.300 |
| Right AF<sup>4</sup> | 0.45 (0.04) | 0.45 (0.04) | t(71)= -0.76 | 0.452 |
| Left UF<sup>5</sup> | 0.25 (0.03) | 0.27 (0.04) | t(71)= -2.39 | **0.020** |
| Right UF<sup>5</sup> | 0.28 (0.03) | 0.29 (0.04) | t(71)= -1.91 | 0.060 |

<sup>1</sup>Child Behavior Checklist; <sup>2</sup>Fractional anisotropy; <sup>3</sup>Anterior thalamic radiation; <sup>4</sup>Arcuate fasciculus; <sup>5</sup>Uncinate fasciculus

### Associations of internalizing problems and mean-tract FA of cerebral tracts

Table 2 shows the results of multiple regression models for the associations of internalizing problems and mean-tract FA of bilateral AF and UF. Supplementary Table 1 shows the results of multiple regression models for bilateral ATR, none of which were significant.

**Table 2.** Regression analysis of internalizing problems and mean-tract FA of the Arcuate Fasciculus and Uncinate Fasciculus.

| Model | 1 | 2a | 2b | 3a | 3c | 4a | 4b | 5a | 5b |
| --- | --- | --- | --- | --- | --- | --- | --- | --- | --- |
| Birth Group | 3.07 | 2.69 | -83.32* | 3.48 | -60.22* | 2.93 | -0.04 | 2.05 | -56.73* |
| Sex | -1.33 | -1.25 | -1.82 | -1.04 | -2.34 | -1.17 | -1.15 | -1.29 | 1.52 |
| Left AF<sup>1</sup> |  | -47.03 | **-134.75\*\*** |  |  |  |  |  |  |
| Left AF<sup>1</sup> x birth group |  |  | **182.38\*** |  |  |  |  |  |  |
| Right AF<sup>1</sup> |  |  |  | -51.07 | **-132.52\*\*** |  |  |  |  |
| Right AF<sup>1</sup> x birth group |  |  |  |  | **141.03\*** |  |  |  |  |
| Left UF<sup>2</sup> |  |  |  |  |  | 8.78 | 1.24 |  |  |
| Left UF<sup>2</sup> x birth group |  |  |  |  |  |  | 11.77 |  |  |
| Right UF<sup>2</sup> |  |  |  |  |  |  |  | 8.42 | **-127.28<sup>a</sup>** |
| Right UF<sup>2</sup> x birth group |  |  |  |  |  |  |  |  | **212.47\*** |
| R2 | 0.03 | 0.05 | **0.14\*** | 0.07 | **0.13\*** | 0.03 | 0.03 | 0.03 | **0.12<sup>a</sup>** |
| R2 change | - | 0.02 | **0.09\*** | 0.04 | **0.06\*** | 0.00 | 0.00 | 0.00 | **0.08\*** |

Note: \* $p < 0.05$ ; \*\* $p < 0.01$ ; <sup>a</sup> $p < 0.10$ ; <sup>1</sup>Arcuate fasciculus; <sup>2</sup>Uncinate fasciculus. R2 change values reflect an increase in variance accounted for in reference to the model with covariates (model 1). For models with interaction terms, the R2 change values are in reference to the model which examined the main effects adjusting for sex and birth group. Significant values are in bold.

Model 1 in Table 2 shows that birth group and sex did not make a significant contribution to internalizing problem T-scores. Model 2a and 2b summarize the analyses of left AF. Model 2a shows that mean-tract FA of left AF was not associated with internalizing problem T-score as a main effect. Model 2b, which included group-by-tract interaction, significantly increased the overall model fit by 9% of variance (*p*=0.011), suggesting that the degree of association between mean-tract FA of left AF and internalizing problem T-scores differed by birth group. Figure 1A shows that lower mean-tract FA of left AF was associated with higher internalizing problem T score only in children born at term, but not children born preterm (b_left AF Term_=-134.75, *p*_left AF Term_ =0.010, b_left AF preterm_=47.64, *p*_left AF preterm_ =0.340).

**Figure 1:**
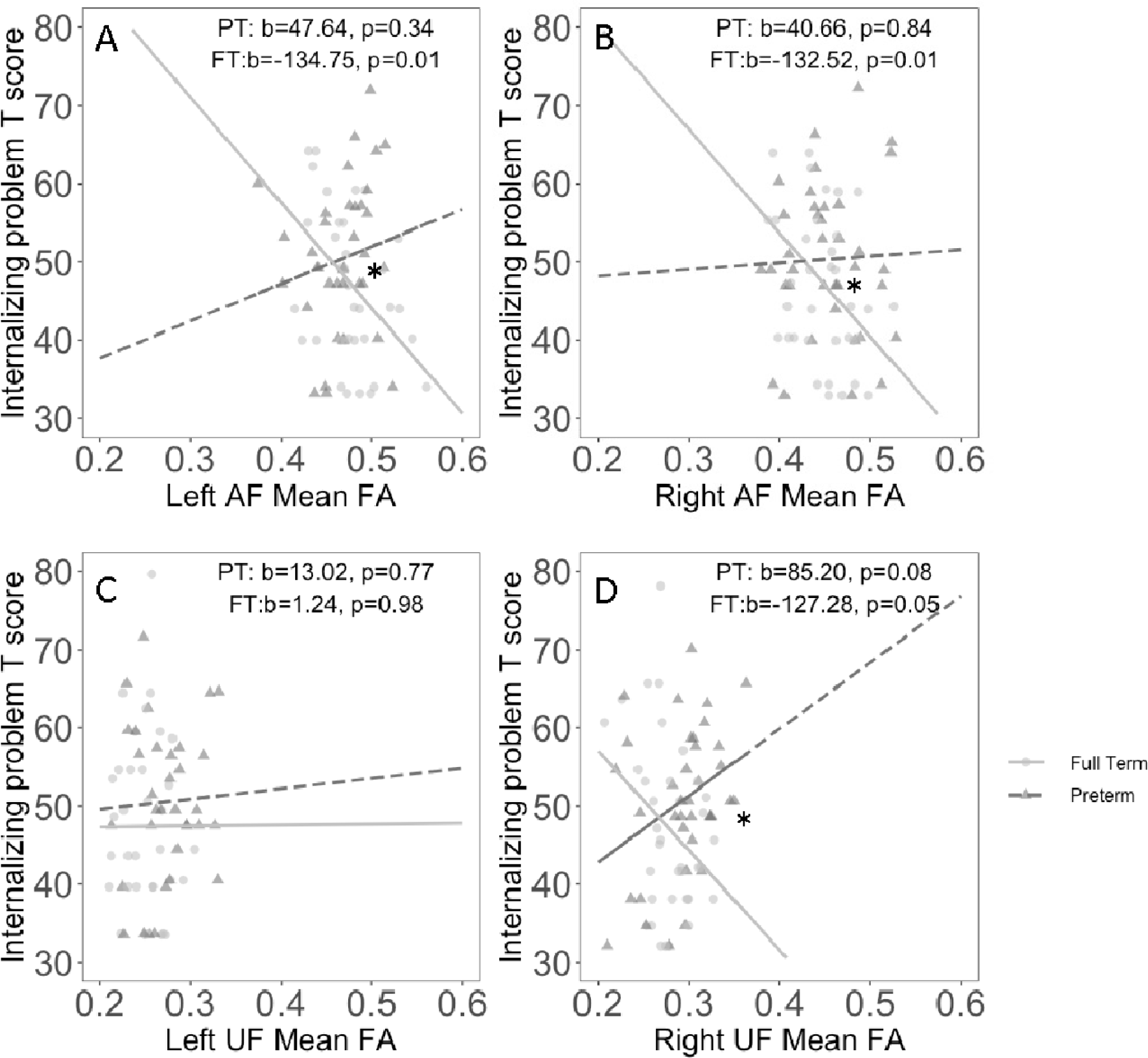
Association between mean-tract FA and internalizing problem T-score. Subplots (A, B) shows that lower mean-tract FA of left or right AF was associated with higher internalizing problem T-score in children born at term; there was no association between mean-tract FA of AF and internalizing problem T-score in children born preterm. Subplot C shows that there was no association between mean-tract FA of left UF and internalizing problem T-score in either birth group. Subplot D shows that while interaction term is significant, there was negative association between the mean-tract FA of right UF and internalizing problems in children born at term, and marginal positive association between the mean-tract FA of right UF and internalizing problems in children born preterm. *statistically significant interaction term.

Model 3a and 3b summarize the analyses of right AF. Model 3a shows that mean-tract FA of right AF was not associated with internalizing problem T-score as a main effect. Model 3b, which included the group-by-tract interaction term significantly increased the model fit by 6% of variance (*p=*0.030), suggesting that the degree of association between mean-tract FA of right AF and internalizing problem T-scores differed by birth group. Figure 1B again shows that lower mean-tract FA of right AF was associated with higher internalizing problem T score only in children born at term, but not children born preterm (b_right AF Term_=-132.52, *p*_right AF Term_ =0.010, b_right AF preterm_=8.50, *p*_right AF preterm_ =0.840).

Model 4a and 4b summarize the analyses of left UF. These models were not statistically significant (Figure 1C).

Model 5a and 5b summarize the analyses of right UF. Model 5a shows that the mean-tract FA of right UF was not associated with internalizing problem T-score as a main effect. Model 5b, which included the group-by-tract interaction significantly increased the model fit by 12% of the variance (*p*=0.014), suggesting that the degree of association between mean-tract FA of right UF and internalizing problem T-scores differed by birth group. Figure 1D shows the significant negative association between internalizing problems and mean FA of right UF in children born at term, and a trending positive association between internalizing problem and mean FA of right UF in children born preterm (b_right UF Term_=-127.28, *p*_right UF Term_ =0.050, b_right UF preterm_=85.20, *p*_right UF preterm_ =0.080).

### Associations of externalizing problems and mean-tract FA of cerebral tracts

We then investigated the associations of externalizing problems and mean-tract FA of cerebral tracts. Table 3 shows the results of multiple regression models for bilateral AF and UF, and Supplementary Table 2 shows the results of multiple regression models for bilateral ATR, none of which were statistically significant.

**Table 3.**
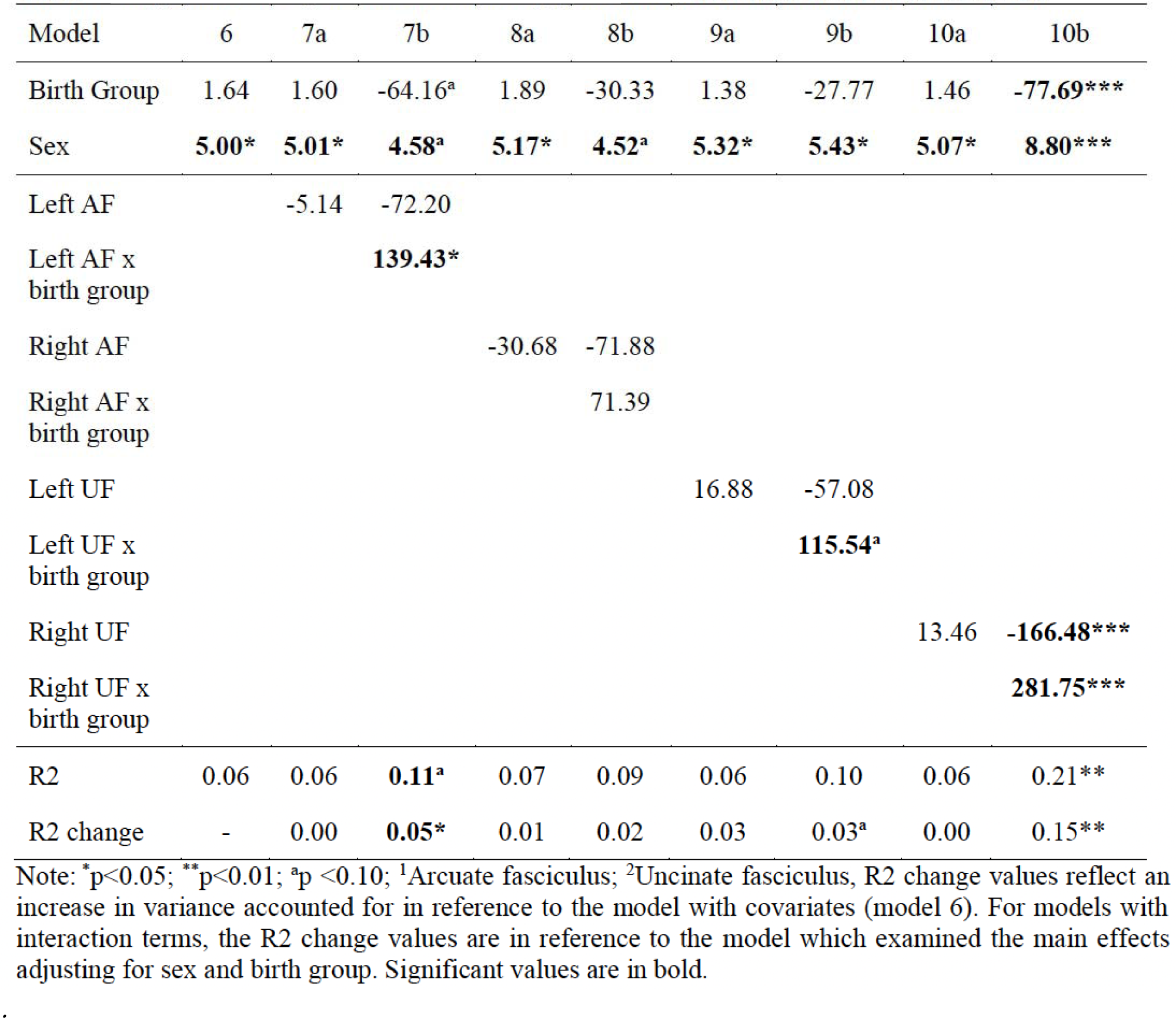
Regression analysis of externalizing problems and mean-tract FA of AF or UF.

Model 6 shows that while sex significantly contributes as a variable, the overall model was not statistically associated with externalizing problem T-scores, accounting for 6% of the variance. Model 7a demonstrates that mean-tract FA of left AF was not significantly associated with externalizing problem T-scores after controlling for sex and birth group. Model 7b, which included the group-by-tract interaction significantly increased the model fit by 5% of accounted variance (*p*=0.048), implying that a difference in degree of association between the mean-tract FA of left AF and externalizing problems by birth group (Figure 2A). The overall model approached statistical significance (*p*=0.087); however, the slope for each birth group was not statistically significant (b_left AF Term_=-72.20, *p*_left AF Term_ =0.140, b_left AF preterm_=67.23, *p*_left AF preterm_ =0.180).

**Figure 2:**
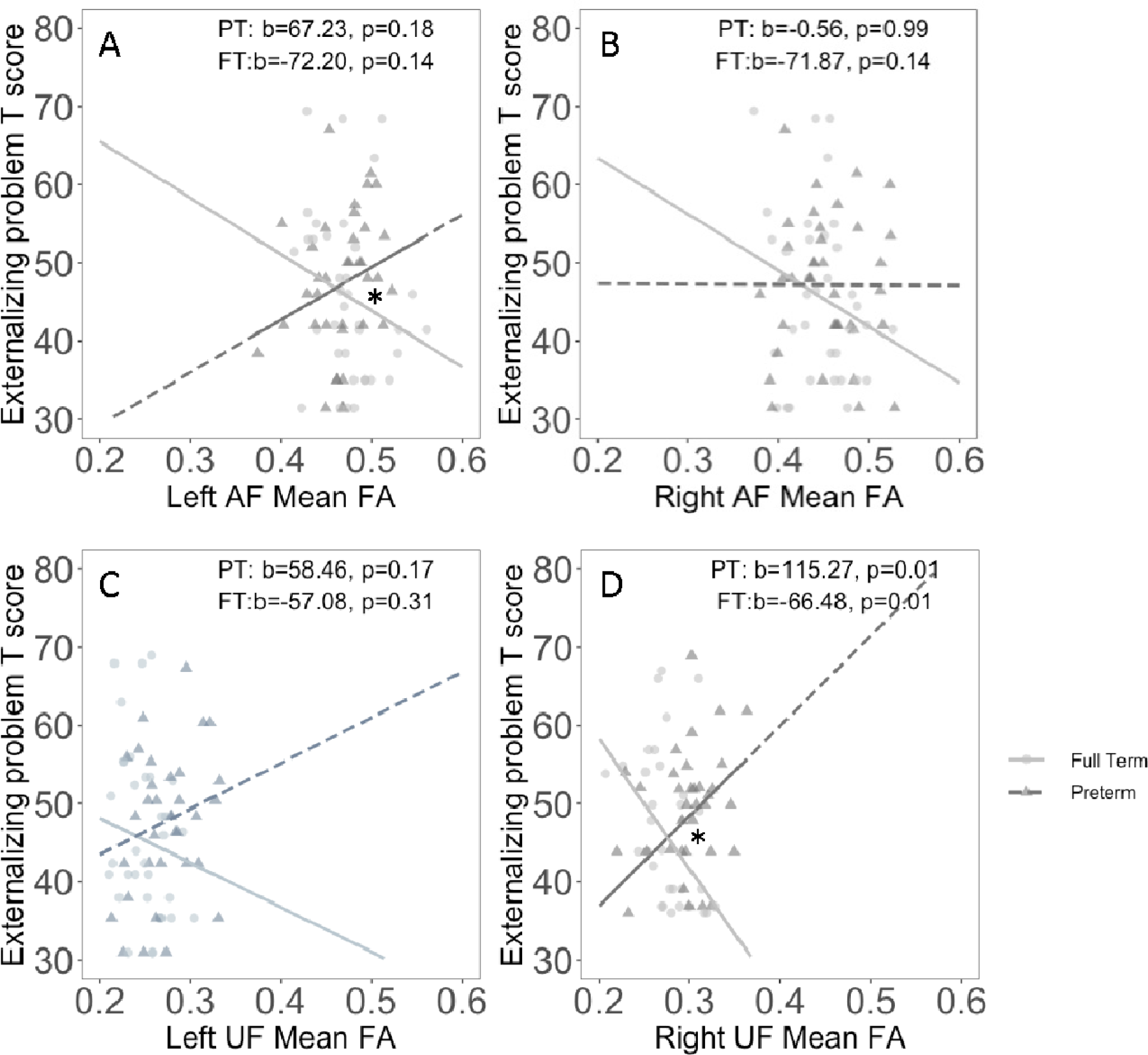
Association between mean-tract FA and externalizing problem T-score. Subplot A shows that while interaction between the mean-tract FA of left AF and externalizing problem T-score by group is significant, there was no association between mean-tract FA of left AF and externalizing problem T-score for each group. Subplot B and C show no association between mean-tract FA of right AF and left UF and externalizing problem T-score in either birth group, respectively. Subplot D shows the negative association between the mean-tract FA of right UF and externalizing problems in children born at term, and positive association between the mean-tract FA of right UF and externalizing problems in children born preterm when adjusting for sex and birth group. * statistically significant interaction term.

Model 8a and 8b summarize the analyses of right AF. These models were not statistically significant (Figure 2B). Model 9a and 9b summarize the analyses of the left UF. These models also were also not statistically significant (Figure 2C).

Model 10a and 10b summarize the analyses of right UF. Model 10a shows no statistical association between mean-tract FA of right UF and externalizing problem T-score as a main effect, after controlling for covariates. Model 10b, which includes group-by-tract interaction term shows that the overall model was statistically significant(*p*=0.002), and the interaction term increased model fit by 15% of variance (*p*=0.001), implying the associations differed by birth group. Figure 2D depicts that lower mean-tract FA of right UF was associated with higher externalizing T-scores in children born at term, and lower mean-tract FA of right UF was associated with lower externalizing T-scores in children born preterm. (b_right UF Term_=-166.58, *p*_right UF Term_ =0.01, b_right UF preterm_=115.27, *p*_right UF preterm_ =0.01).

To determine if the association of right UF and internalizing problems was driven by the shared variance of internalizing and externalizing problems, we first ran a regression model that assessed contribution of the mean-tract FA and group-by-tract interaction of right UF to externalizing problems while controlling for internalizing problem T-score, sex and birth group. The overall model was statistically significant (*p*<0.001), and the interaction term of the right UF mean-tract FA x birth group continued to be significant (*p*=0.010). The negative association among children born at term and positive association among children born preterm continued to be statistically significant (b_right UF Term_=-114.11, *p*_right UF Term_ =0.04, b_right UF preterm_=80.22, *p*_right UF preterm_ =0.05) (Supplementary Figure 1). This finding suggests that the right UF is associated with both internalizing and externalizing problems and not solely with the shared variance between them.

## Discussion

In summary, the aim of this study was to investigate whether microstructure of three white matter circuits connecting to the frontal cortex would be associated with internalizing or externalizing behavioral problems in school-aged children, and if the association would differ between children born at term and children born preterm. We found that mean-tract FA of the right and left AF and of the right UF was negatively associated with internalizing problems only in children born at term. Mean-tract FA of right UF was also negatively associated with externalizing problems in children born at term, and positively associated with externalizing problems in children born at term.

In this sample, mean T-scores on the CBCL/6-18 for both children born at term and preterm were close to 50, the mean T-score for the general population. Higher scores on this scale represent greater number and/or severity of behavioral problems and are therefore less favorable. In the children born at term, higher scores on the CBCL/6-18 scales were associated with lower mean-tract FA. These results suggest that within this term group, the factors associated with increasing FA, such as higher fiber density and/or increased myelination, were associated with a favorable behavioral profile. By contrast, the association of externalizing behavioral problems and FA of the right UF was positive in the preterm group, meaning that children with higher number and/or severity of behavioral problems had higher FA. Higher fiber density and/or increased myelination in the preterm group were associated with a less favorable behavioral profile. Different patterns of association between cognitive measures and FA in children born at term and preterm have been found in this sample of children (Bruckert et al., 2019; Dodson et al., 2018) and in other samples (Kelly et al., 2023; Travis et al., 2015). The different pattern of association suggests that preterm birth, with its associated perinatal injury to oligodendrocyte precursors and downstream effects of white matter circuits, results in distinctive brain-behavior associations.

The results we found differed from those reported by Loe et al (Loe et al., 2013), in which associations were found exclusively among children born preterm and in the opposite direction. We presume that the reason is the difference in the analytic methods of white matter characterization. That study relied on Tract-Based Spatial Statistics whereas we used probabilistic tractography, a presumably more accurate method. It is difficult to interpret which white matter properties may set up for findings of positive associations of externalizing behavioral problems with FA. FA is a summary variable of multiple, interacting microstructural features; white matter tracts have crossing fibers that complicate the interpretation of FA (Figley et al., 2022). One experimental strategy to learn about the microstructural features causing the positive association in the preterm group would be to use convergence of different advanced MRI methods, each sensitive to a distinct microstructural feature. For now, we can only say that the two birth groups differ in the underlying neurobiology of behavioral problems.

### Internalizing problems and the AF

In the children born at term, we found that internalizing problems were negatively associated with bilateral AF. The AF connects the frontal, parietal and temporal cortex. Emerging evidence suggests that the AF participates in attention and self-regulation. AF, together with superior longitudinal fasciculus-III branch and inferior frontal occipital fasciculus, have been labeled a “ventral attention network” (Hattori et al., 2018; Urger et al., 2015). AF is also an important tract for language processing. Specifically, many studies find that the left AF is associated with reading and language skills (Wandell and Yeatman, 2013) and language-based working memory (Barbeau et al., 2023). It is notable that internalizing behavioral problems are commonly seen in children with attentional weaknesses, including Attention Deficit Hyperactivity Disorder and with language and/or reading problems (Bishop et al., 2019; Donolato et al., 2022; Peterson et al., 2017). One possibility is that the fibers within the AF, associated with internalizing problems, are the identical fibers engaged in transfer of information for language and reading. That would mean that the signals in both systems travel the same circuits. Alternatively, the AF may have distinct subsets of fibers that serve to transfer information about behavioral issues, attention, or language and reading. With the methods we used in this study, we could not dissect the AF with sufficient detail to determine if the same fibers would be associated with internalizing problems, attention, and reading. Future studies using different methods may be able to investigate the AF in greater detail than we could.

### Behavioral problems and the UF

We found the negative association of externalizing problems and right UF in children born at term, and positive association in children born preterm. We also found the negative association of internalizing problems and right UF in children born at term and approaching positive association in children born preterm. The association between externalizing problems and right UF did not change when we controlled for scores on the scales for internalizing problems.

The UF is an association white matter tract that connects the temporal lobe with prefrontal regions associated with emotional regulation, impulsivity, and high-order cognitive skills (Andre et al., 2020; Travis et al., 2015; Von Der Heide et al., 2013). Many studies suggest that structural and functional connectivity of the UF plays a role in linking the generation of emotions (from within the limbic system) to self-regulation (controlled by the frontal cortex). For example, in children aged 9-16, diffusion-tensor imaging and functional MRI demonstrated negative associations between UF FA and activation of the amygdala to pictures of sad and happy faces, though in that study there were no significant associations to frontal cortex (Swartz et al., 2014). In a study that evaluated white matter characteristics in children with and without autism, higher FA in the UF was associated with better self-regulation in children without autism, whereas higher FA was associated with worsening self-regulation in children with autism (Ni et al., 2020).

The association between mean-tract FA of the right UF and behavioral problems was negative in children born at term, meaning that lower T-score (indicative of fewer or less severe behavioral problems) was associated with higher FA. In this age group, behavioral problems (such as aggression or anxiety) are commonly seen in children with poor self-regulation, such as within children with impulsivity or ADHD (Reimherr et al., 2017). We were uncertain whether the same UF fibers might be serving to transfer information regarding both internalizing and externalizing behavioral characteristics and the associations were based on shared variance between the internalizing and externalizing T-scores. For that reason, we ran regression models for externalizing behavioral problems, including the scores on the internalizing problem scales. We found that the degree of association did not change substantially, suggesting that the UF is associated with both internalizing problems and with externalizing problems independently, not based on the shared variance between the scales. Again, with our methods, we could not dissect UF into specific fibers and investigate the role of specific fibers to specific behavioral problems (internalizing problems, externalizing problems). We suggest future studies investigate UF function and connectivity in school-aged children using methods that might allow precision in the dissection of the tract.

Among children born preterm, we found that the association between mean-tract FA of the right UF and behavioral problems was positive, implicating higher behavioral problems with higher FA. The association of externalizing behavioral problems and mean-tract UF was not fully explained by the scores on the internalizing problem scale. The gray and white matter of children born preterm undergo altered white matter integrity (Duerden et al., 2019). One possible explanation is that UF has specific or independent roles in internalizing problems and in externalizing problems. The complex white matter injury secondary to preterm birth and subsequent development of white matter circuits changed the direction of association. We acknowledge that previous literature demonstrates inconsistent results among children born preterm. For example, in Kanel’s study, the term-equivalent mean-tract FA of right UF in children born preterm was associated with higher “emotion moderation”, but not among children born at term (Kanel et al., 2021).

In Gilchrist et al’s study that examined association between three different fiber-specific measures and concurrent behavior in 7-year-olds with and without history of preterm birth, bilateral UF metrics were associated with internalizing problems and externalizing problems regardless of birth group (Gilchrist et al., 2023). Among the 13-year olds, the only significant associations were between fiber density of the right UF and both internalizing and externalizing problems in the term group. Different findings may relate to the methods used to characterize white matter and to differences in the study population. However, these findings further support a role for the UF in behavior problems. We suggest future studies in large samples using multiple advanced MRI methods to investigate the role of UF in behavioral problems in children born preterm.

### Behavioral problems and ATR

In this study, we did not see association between behavioral problems and ATR in either birth group. In Gilchrist et al (Gilchrist et al., 2023) there was no association between internalizing or externalizing problems and fiber density, a measurement of specific fiber population density within a voxel, of the ATR. However, they found association between internalizing and externalizing problems and fiber-bundle cross section and fiber-density-and-cross-section, measures that relate to transferring information across white matter. In Loe’s study, they did not find association between internalizing problems and ATR in either birth group but found association between attention problems and ATR in children born at term (Loe et al., 2013). Future studies should verify the contribution of ATR in children.

### Preterm birth

Among children born preterm, we did not identify an association between internalizing or externalizing behavioral problems and mean-tract FA in any tract except for the right UF. Among children born less than 32 weeks of gestational age, diffuse white matter injury is commonly seen (Schneider and Miller, 2019). The diffuse white matter injury leads to reactive astrogenesis and gliosis (Schneider and Miller, 2019), which results in altered white matter integrity and functional connectivity within and between hemispheres (Duerden et al., 2019). The white matter integrity varies across birth group (Travis et al., 2015). In children born preterm, the overall behavioral characteristics are perhaps due to a sum of diffuse atypical white matter difference rather than a white matter difference in selected cerebral tracts.

### Limitations and future directions

Our study has several limitations. All participants were recruited within the San Francisco Bay Area, and the socioeconomic status of the sample was high. The results may not generalize to a sample of low socioeconomic status. Moreover, the collected behavioral measurements were derived from parent-reported Child Behavior Checklist; parent-reported scores for internalizing problems may not have fully reflected the participants’ internal feelings of anxiety or depression. Future studies should consider using specific self-reported instruments as well as parent reports. The children in this study had average scores on the behavioral problem scales and there were no differences between the birth groups. Future studies might recruit children with higher behavioral problem scores than the children we studied here.

Our study found significant associations between internalizing and externalizing behavioral problems and white matter metrics of cerebral circuits connecting to the frontal cortex. We noted that mean-tract FA explained a modest proportion of the variance in behavioral problem scores, among the children born at term. We think that the limited strength of the association is because behavioral problems are caused by a convergence of biological and environmental factors (Hatoum et al., 2018b). The environment may play an etiological role, such that children from stressful environments are more likely to develop behavioral problems than children from easy and manageable environments. In addition, environmental factors are critical for interpretation of what is and what is not a behavioral problem. Often it is not the behavior *per se* that is the issue, but the behavior within a social context. For example, quiet withdrawal, an internalizing problem, may be appropriate in the context of a religious service or within a classroom setting, though inappropriate at a birthday party. Children with internalizing or externalizing behavioral problems may be able to demonstrate behavioral control within environmental contexts that cultivate their motivation to remain in control.

We recognize that we are unable to make any claims about causality; we do not know if the presence of behavioral problems influences white matter microstructure, or, conversely if features of white matter microstructure lead to the onset of behavioral problems. A longitudinal study investigating the causal relationship of anxiety and white matter integrity in anxious preadolescent girls showed that higher anxiety led to decreased whole-brain white matter integrity, but without concurrent association (Aggarwal et al., 2022). In children aged 6-10 years, presence of internalizing problems and externalizing problems predicted slow growth in whole-brain white matter integrity, but not vice versa (Muetzel et al., 2018). Additional research should investigate longitudinal associations between the behavioral characteristics and white matter metrics.

If further studies replicate the associations of behavioral problems and white matter microstructure of the AF and UF, the findings may have clinical implications. The first-line treatment for children with clinically significant internalizing problems, such as anxiety and depression, is cognitive behavioral therapy. Cognitive behavioral therapy nurtures self-regulation and control through skills taught verbally. White matter microstructure of the AF and UF may predict response to cognitive behavior therapy or may change after treatment with cognitive behavior therapy, representing adjustments in the flow of information in the brain. Future studies may consider investigating longitudinal change in the association of behavioral problems and white matter integrity over the course of development and in response to evidence-based therapy.

## Data Availability

All data produced in the present study are available upon reasonable request to the authors

## Acknowledgments

This work was supported by the National Institutes of Health (NICHD grant RO1-HD069162 A and 2RO1-HD069150) and Health Resources and Services Administration Maternal Child Health Bureau (T77MC09796) to Heidi M Feldman.

## Supplementary Materials

**Supplementaiy Table 1.** Internalizing problem and mean-tract FA of ATR.

| Model | 1 | 11a | 11b | 12a | 12b |
| --- | --- | --- | --- | --- | --- |
| Birth Group | 3.07 | 2.42 | -22.42 | 2.32 | -5.49 |
| Sex | -1.33 | -1.30 | -1.50 | -1.44 | -1.51 |
| Left ATR<sup>1</sup> |  | -38.09 | -62.95 |  |  |
| Left ATR<sup>1</sup> x birth group |  |  | 56.20 |  |  |
| Right ATR<sup>1</sup> |  |  |  | -43.06 | -49.50 |
| Right ATR<sup>1</sup> x birth group |  |  |  |  | 18.43 |
| R2 | 0.03 | 0.05 | 0.06 | 0.06 | 0.07 |
| R2 change | - | 0.02 | 0.01 | 0.03 | 0.01 |

Note: \*p<0.05; \*\*p<0.01; <sup>a</sup>p <0.10; <sup>1</sup>Anterior thalamic radiation. R2 change values reflect an increase in variance accounted for in reference to the model with covariates (model 1). For models with interaction terms, the R2 change values are in reference to the model which examined the main effects of adjusting for sex and birth group. Significant values are in bold.

**Supplementaiy Table 2.**
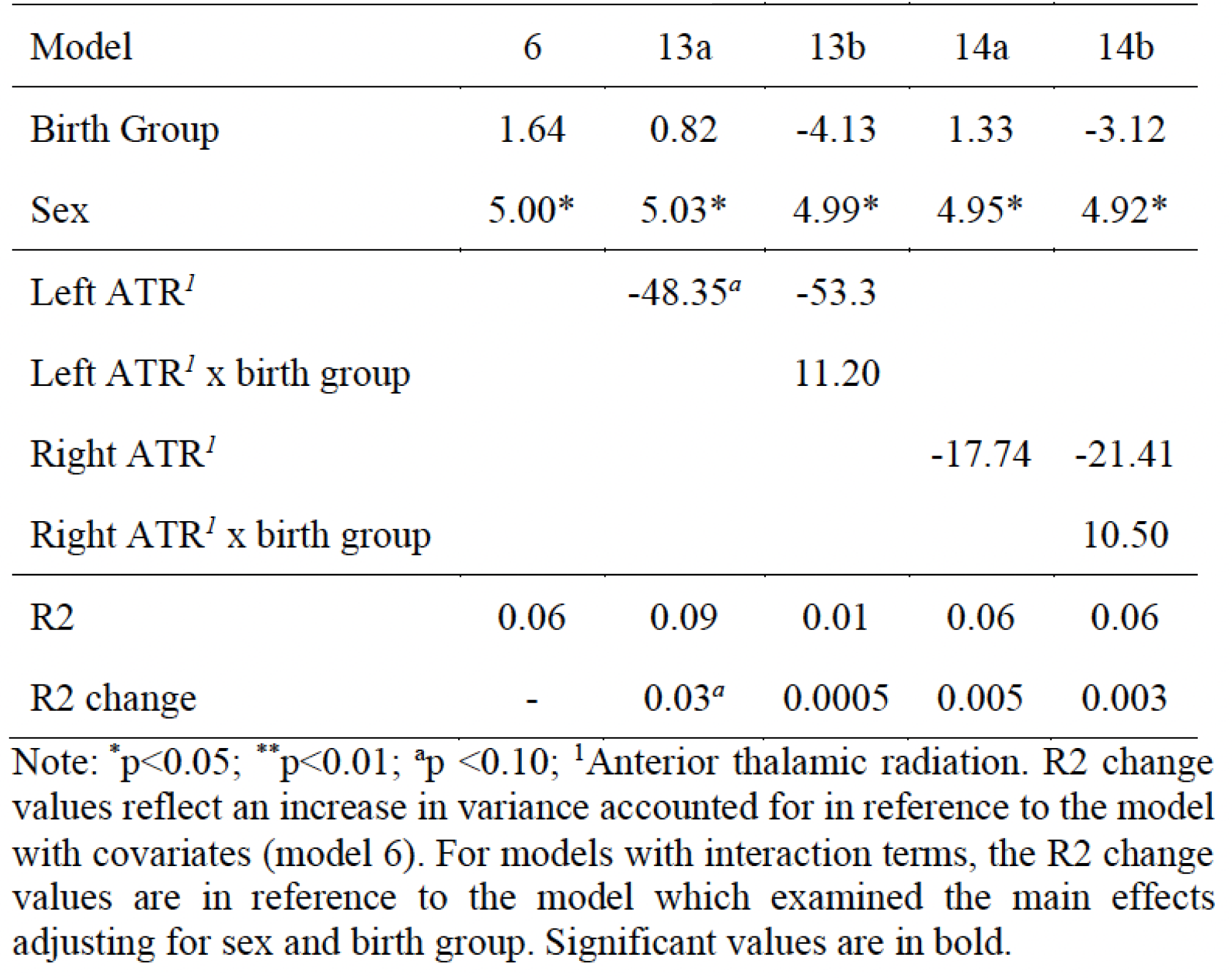
Externalizing problem and mean-tract FA of ATR.

| Model | 6 | 13a | 13b | 14a | 14b |
| --- | --- | --- | --- | --- | --- |
| Birth Group | 1.64 | 0.82 | -4.13 | 1.33 | -3.12 |
| Sex | 5.00* | 5.03* | 4.99* | 4.95* | 4.92* |
| Left ATR<sup>1</sup> |  | -48.35<sup>a</sup> | -53.3 |  |  |
| Left ATR<sup>1</sup> x birth group |  |  | 11.20 |  |  |
| Right ATR<sup>1</sup> |  |  |  | -17.74 | -21.41 |
| Right ATR<sup>1</sup> x birth group |  |  |  |  | 10.50 |
| R2 | 0.06 | 0.09 | 0.01 | 0.06 | 0.06 |
| R2 change | - | 0.03<sup>a</sup> | 0.0005 | 0.005 | 0.003 |

Note: \*p<0.05; \*\*p<0.01; <sup>a</sup>p <0.10; <sup>1</sup>Anterior thalamic radiation. R2 change values reflect an increase in variance accounted for in reference to the model with covariates (model 6). For models with interaction terms, the R2 change values are in reference to the model which examined the main effects adjusting for sex and birth group. Significant values are in bold.

**Figure S1.**
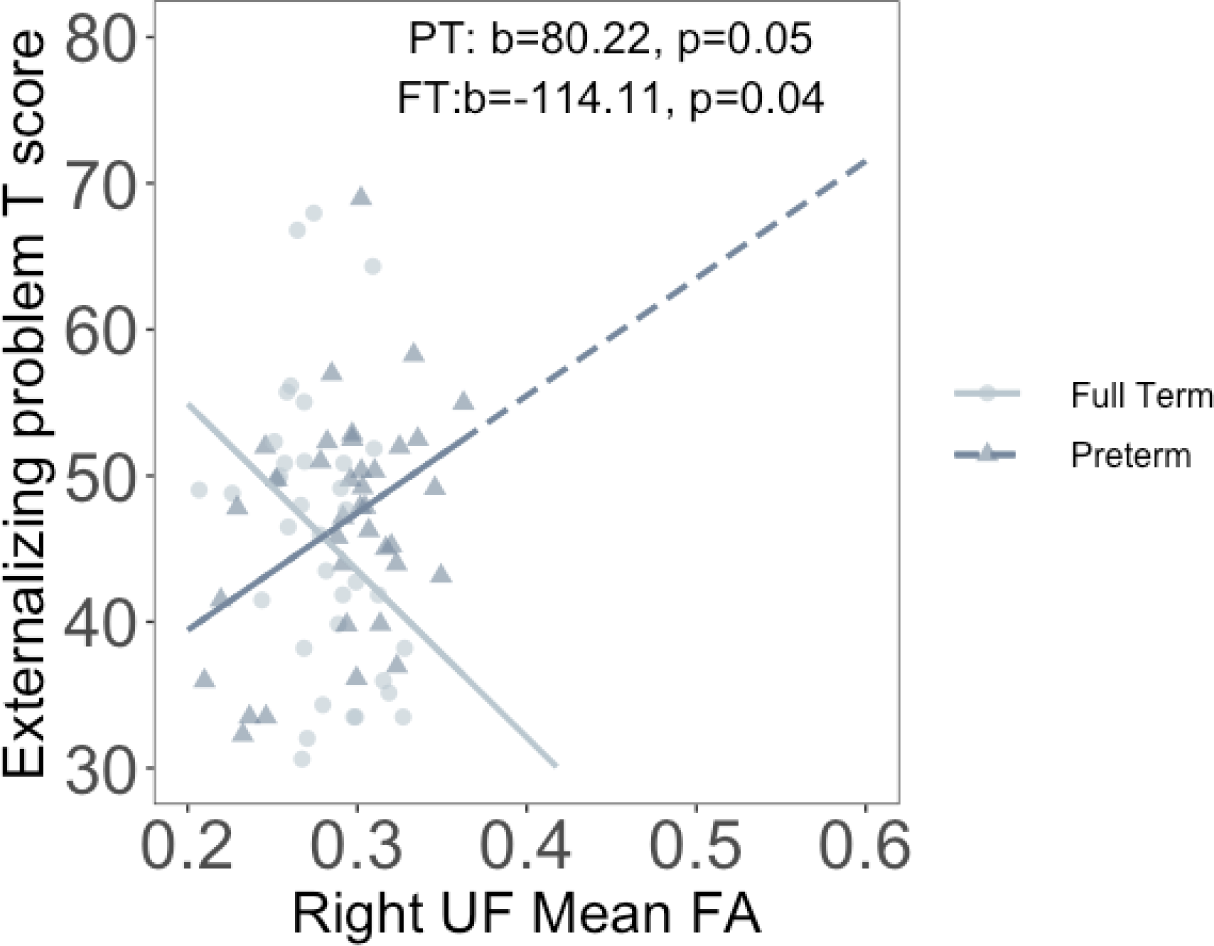
Association between mean-tract FA of right UF and externalizing problem T-score by birth group, after controlling for internalizing problem T-score, sex and birth group. There continues to be a negative association between mean-tract FA of right UF and externalizing problems in children born at term. There is a marginal positive association between mean-tract FA of right UF and externalizing problems in children born preterm.

